# Progression of prostate cancer reprograms MYC-mediated lipid metabolism via lysine methyltransferase 2A

**DOI:** 10.1101/2022.06.04.22276001

**Authors:** Nichelle C. Whitlock, Margaret E. White, Brian J. Capaldo, Anson T. Ku, Supreet Agarwal, Lei Fang, Scott Wilkinson, Shana Y. Trostel, Zhen-Dan Shi, Falguni Basuli, Karen Wong, Elaine M. Jagoda, Kathleen Kelly, Peter L. Choyke, Adam G. Sowalsky

## Abstract

**Background:** The activities of MYC, the androgen receptor, and its associated pioneer factors demonstrate substantial reprogramming between early and advanced prostate cancer. Although previous studies have shown a shift in cellular metabolic requirements associated with prostate cancer progression, the epigenetic regulation of these processes is incompletely described. Here, we have integrated chromatin immunoprecipitation sequencing (ChIP-seq) and whole-transcriptome sequencing to identify novel regulators of metabolism in advanced prostate tumors characterized by elevated MYC activity.

**Results:** Using ChIP-seq against MYC, HOXB13, and AR in LNCaP cells, we observed redistribution of co-bound sites suggestive of differential KMT2A activity as a function of *MYC* expression. In a cohort of 177 laser-capture microdissected foci of prostate tumors, *KMT2A* expression was positively correlated with MYC activity, AR activity, and *HOXB13* expression, but decreased with tumor grade severity. However, *KMT2A* expression was negatively correlated with these factors in 25 LuCaP patient-derived xenograft models of advanced prostate cancer and 99 laser-capture microdissected foci of metastatic castration-resistant prostate cancer. Stratified by *KMT2A* expression, ChIP-seq against AR and HOXB13 in 15 LuCaP patient-derived xenografts showed an inverse association with sites involving genes implicated in lipid metabolism, including the arachidonic acid metabolic enzyme *PLA2G4F*. LuCaP patient-derived xenograft models grown as organoids recapitulated the inverse association between *KMT2A* expression and fluorine-18 labeled arachidonic acid uptake *in vitro*.

**Conclusions:** Our study demonstrates that the epigenetic activity of transcription factor oncogenes exhibits a shift during prostate cancer progression with distinctive phenotypic effects on metabolism. These epigenetically driven changes in lipid metabolism may serve as novel targets for the development of novel imaging agents and therapeutics.

## BACKGROUND

Prostate cancers (PCa) are distinctively sensitive to the transcriptional activity of the androgen receptor (AR) during tumorigenesis and in response to hormone-based therapies for advanced disease [1]. In early PCa, the AR’s role in mediating luminal cell terminal differentiation undergoes a distinctive switch as AR remains active in progressively less-differentiated disease [2]. Concurrent with early transformation of these cells, the *MYC* proto-oncogene reactivates and opposes the antiproliferative functions of AR to drive continued cell growth [3, 4]. Although elevated MYC protein levels are detectable very early in both pre-neoplastic and neoplastic prostate luminal cells, unlike many other tumor types, its mRNA expression remains largely uncoupled from genomic alterations affecting the *MYC* locus with aneuploidies involving most of chromosome 8q [5–7]. By contrast, in advanced and metastatic castration-resistant prostate cancer (mCRPC), *MYC* frequently undergoes high-level or focal amplification which significantly co-occurs with focal gains of the *AR* gene body and/or its enhancer [8, 9].

Although increased transcriptional output is canonically associated with increased expression of transcription factors, both MYC and AR show evidence of rewiring or reprogramming upon progression of primary PCa to advanced disease, especially in the context of hormonal therapies used for treating recurrent PCa [10]. Hormone- and context-dependent activities of these master regulators further involve pioneer factors that mediate AR-driven transcriptional programming; GATA2, FOXA1, and HOXB13 drive lineage-specific gene expression that is critical for tumor development [11]. Although somatic mutations to *FOXA1* and inherited mutations to *HOXB13* are potential drivers, studies of prostate tumors suggest mostly that differential activity of these factors is associated with disease progression [11]. Recently, the mixed-lineage leukemia (MLL) protein complex, more commonly implicated as a driver of MLL fusion-positive leukemias, was also shown to interact with AR signaling in advanced PCa, demonstrating the increasing complexity of characterizing transcriptional drivers of PCa [12]. *KMT2A* encodes MLL1, a SET domain encoding histone lysine methyltransferase and although it rarely exhibits somatic genomic alterations in PCa, changes to its expression and recruitment to gene promoters and enhancers profoundly impact target gene expression [13].

A primary physiological function of a prostate luminal cell is to secrete prostatic fluid, a major component of which is citrate [14, 15]. This distinctive metabolic requirement for generating citrate precursors is maintained during PCa tumorigenesis with elevated flux through the tricarboxylic acid cycle despite tumorigenic hypoxic conditions [15]. In addition, MYC and AR cooperate to facilitate *de novo* synthesis of polyunsaturated long-chain fatty acids to accommodate increased demand for phospholipids on account of increased cell division events [16–18]. The 20-carbon arachidonic acid (ArA) is routinely incorporated into phospholipids; ArA is normally acquired dietarily but can be released from phospholipids by deacetylating phospholipases A (PLA) for generating eicosanoids and prostaglandins [19–21]. *PLA2G4F* encodes a Group IV cytosolic phospholipase A_2_ (cPLA_2_) with high specificity for ArA [21]. Although cPLA_2_ proteins are ubiquitously expressed, the relationship between cellular ArA requirements and the effects of AR and MYC on PCa progression are not known.

Here, we report that in prostate cancer cells, differential binding of AR and HOXB13 on account of varying MYC levels centers on genes regulated by *KMT2A*/*MLL1*, which in turn is inversely proportional to the expression of *PLA2G4F* in cohorts of advanced, but not localized, PCa. Patient-derived prostate cancer xenografts grown as organoids have increased uptake of ArA in models with increased MYC activity and *PLA2G4F* expression. Our data demonstrate a phenotypic ramification of epigenetic reprogramming that accompanies disease progression, with potential implications for the design of novel therapeutics and imaging agents.

## RESULTS

### AR and HOXB13 co-localize with MYC at genes regulated by KMT2A

Although MYC activity is a well-known driver of prostate tumorigenesis, its relationship with individual genes and molecular processes remains actively investigated, with current efforts focusing on identifying effector genes or co-factors necessary for its positive association with androgen receptor (AR) activity [7, 10]. Using an institutional cohort of 177 laser capture microdissected (LCM) foci of primary prostate cancer (PCa), we examined the range of global gene expression that tracked with *MYC* transcript abundance. While the top 1% of correlating genes were strongly enriched for ribosomal biogenesis genes, *AR* and multiple AR target genes were also represented (Fig. 1A). Given the known role of MYC in mediating the AR-HOXB13 signaling axis, we analyzed a series of chromatin immunoprecipitation sequencing (ChIP-seq) experiments using antibodies against AR, HOXB13, and MYC, where the level of MYC was modulated by *MYC*-targeting hairpins (shMYC) [22, 23]. The overlap between MYC and AR or MYC and HOXB13 binding identified targets that may contribute to its function in primary PCa (outlined in Fig. 1B), which in turn pinpointed potential master regulators that regulate tumorigenesis.

**Figure 1.**
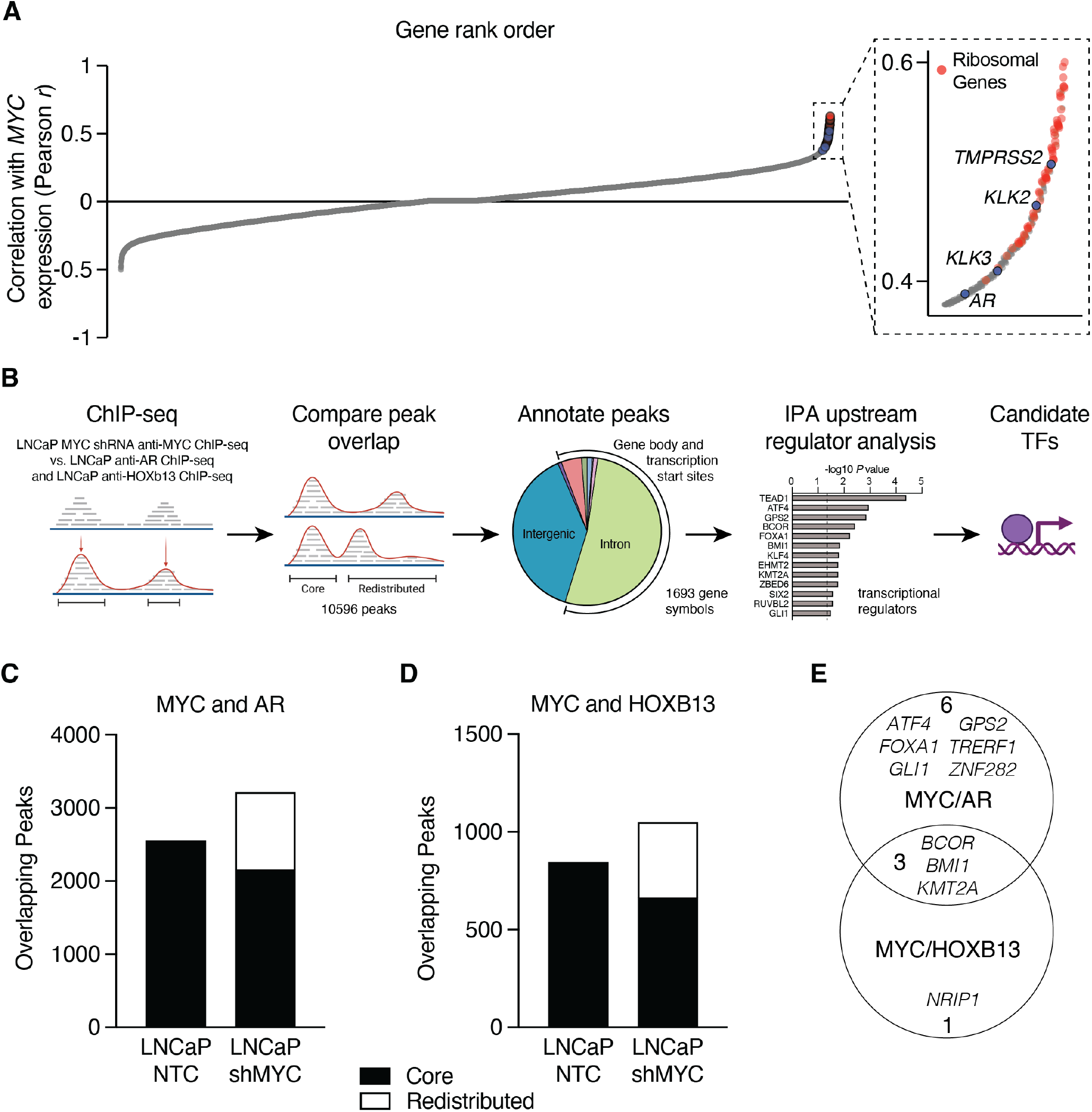
Identification of transcription factors associated with MYC redistribution at AR-and HOXB13-occupied sites. **A**. Ranked order depiction of genes whose expression correlate with *MYC* in an institutional cohort of 177 laser-capture microdissected prostate cancer tumor foci. **B**. Schematic of the strategy used to identify upstream transcription regulators based on co-occupied peaks in LNCaP ChIP-seq data. anti-MYC [22], anti-AR, and anti-HOXB13 ChIP-seq data [23] used for analyses. **C, D**. Bar plots showing the total peak number of MYC co-occupied sites, characterized as “core” or “redistributed” peaks, for anti-AR (C) and anti-HOXB13 (D) ChIP-seq. **E**. Venn diagraph showing the overlap between transcription regulators identified by MYC/AR and MYC/HOXB13 co-occupied genes and nominated Ingenuity Upstream Regulator Analysis.

Comparison of MYC and AR ChIP-seq data showed ~2600 and ~3200 MYC binding sites that colocalized with AR binding sites in LNCaP/NTC and LNCaP/shMYC cells, respectively; of these, 67% of peaks were shared (2154 of 3213 sites; Fig. 1C). For HOXB13 ChIP-seq, ~850 and ~1050 MYC binding sites were co-occupied in LNCaP/NTC and LNCaP/shMYC cells, respectively, with ~63% overlap (663 of 1049 sites, Fig. 1D). Co-bound sites were categorized as being *core* or *redistributed* and annotated to the nearest gene body or transcription start site. Here, we defined *core* peaks as co-occupied binding sites enriched in both *MYC* high- and low-expressing cells, and *redistributed* peaks as co-occupied binding sites enriched in *MYC* low-expressing cells. The corresponding lists of genes were then used as input for Ingenuity Pathway Analysis (IPA) to identify potential upstream transcriptional regulators (Supplementary Table 1).

This upstream regulator analysis identified nine shared transcription regulators among MYC/AR bound sites and four shared transcription regulators among MYC/HOXB13 bound sites (Fig. 1E). Of these, three were also shared by HOXB13 and AR: *BCOR* (BCL6 corepressor), *BMI1* (B lymphoma Mo-MLV insertion region 1), and *KMT2A* (H3K4 lysine methyltransferase 2A) (Fig. 1E). *BCOR* and *BMI1* encode members of the polycomb repressor complex 1 and are involved in cell differentiation; however, only *BMI1* is implicated in PCa [24, 25]. Recently, a positive relationship between MYC and KMT2A has been documented and given that changes in *KMT2A* expression are associated with advanced disease, we selected *KMT2A* for further analysis [13, 26, 27].

### KMT2A is positively associated with MYC activity in primary, but not advanced, PCa

Our ChIP-seq analyses of genetically-modified prostate cancer cell lines suggested that KMT2A activity may positively mediate the tumorigenic function of MYC. To address this potential interaction further, we analyzed whole-transcriptome sequencing (WTS) data from LCM primary prostate tumor foci (*N* = 177). Case-by-case, *KMT2A* expression was positively correlated with single-sample gene set enrichment analysis (ssGSEA) scores for MYC activity (Fig. 2A; *r* = 0.48, 95% C.I. 0.36 to 0.59, *P*_*r*_ < 0.0001). We next asked whether AR activity or *HOXB13* expression was altered in the context of *KMT2A*. We observed a statistically significant and positive correlation between *KMT2A* and the ssGSEA scores for AR activity (Fig. 2B; *r* = 0.50, 95% C.I. 0.38 to 0.60, *P*_*r*_ < 0.0001) and the expression of *HOXB13* (Fig. 2C; *r* = 0.32, 95% C.I. 0.18 to 0.45, *P*_*r*_ < 0.0001). However, when we subdivided these tumor foci by histological aggressivity (*i*.*e*., Gleason pattern, Gp), these positive associations generally decreased in effect size (Fig. 2A-C) from Gp3 (*N* = 48) to Gp4 (*N* = 101) to Gp5 (*N* = 28).

**Figure 2.**
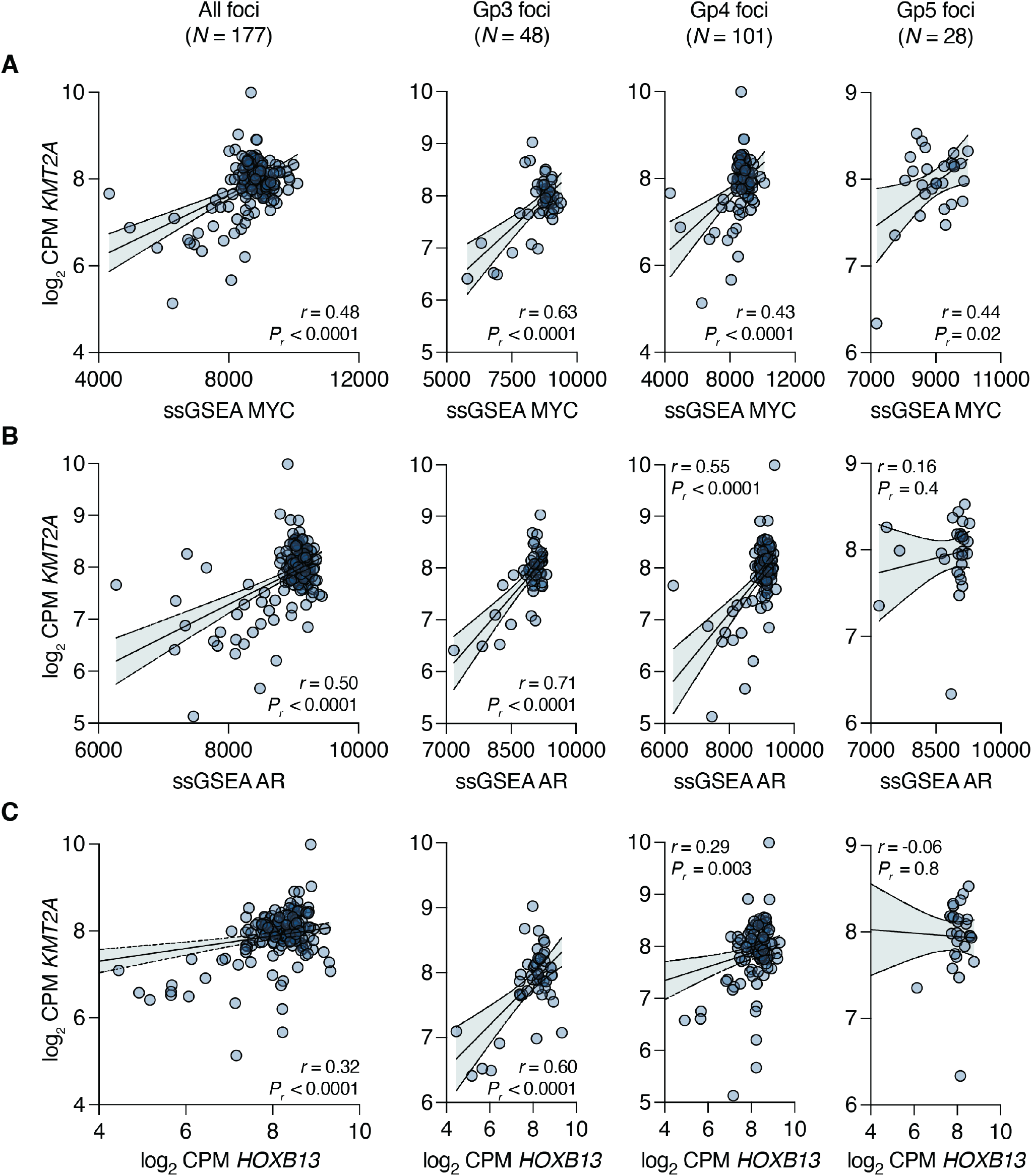
Decreasing association of *KMT2A* expression with prostate cancer drivers during primary disease progression. **A, B, C**. Pearson correlation of the log_2_ CPM expression level for *KMT2A* with a 54-gene ssGSEA MYC activity score (A), a 266-gene ssGSEA AR activity score (B), or the log_2_ CPM expression level for *HOXB13* (C) in a cohort of 177 laser capture microdissected foci of human prostate tumors. The error bars represent the 95% confidence bands for linear regression. For each correlation, the foci are subdivided into Gp3 (left), Gp4 (middle, including intraductal tumors), and Gp5 (right).

To validate this finding, we examined a separate cohort of LCM pairs of adjacent Gp3 and Gp4 tumor foci (from Gleason score 7 PCa) previously analyzed by Affymetrix microarray [28]. Across all foci (*N* = 26), and separately, *KMT2A* expression was positively correlated with MYC activity, AR activity, and *HOXB13* expression (Supplementary Fig. 1). Interestingly however, analysis of WTS data from the TCGA-PRAD cohort (*N* = 484) did not recapitulate these associations (Supplementary Fig. 2), likely due in part to the bulk and admixed nature of TCGA tumor cases.

Nonetheless, these data suggest that at least in a subset of primary PCa, increased expression of *KMT2A* may contribute to tumor development, and that its expression in relationship to MYC activity modulates cancer progression. To assess this relationship in advanced disease, we examined gene expression profiles from WTS of two additional cohorts: a panel of metastatic prostate cancer patient derived xenografts (LuCaP PDX series, *N* = 25, [23, 29–32]) and the West Coast Dream Team-Prostate Cancer Foundation (WCDT-PCF) cohort of metastatic castration-resistant prostate cancer (mCRPC, *N* = 99, [8]). As depicted in Fig. 2A-C, the association between MYC activity, AR activity, *HOXB13* expression and *KMT2A* expression decreased with dedifferentiation associated with higher Gleason patterns. Continuing this trend, we observed no strong association in LuCaP tumors (Fig. 3A), and this association was inversely related in WCDT-PCF tumors (Fig. 3B; *r* = −0.35, 95% C.I. −0.51 to −0.16, *P*_*r*_ = 0.0004). Consistent with this inverse tendency, *KMT2A* expression was also negatively correlated with AR activity and *HOXB13* expression in both the LuCaP PDX (Fig. 3C-D) and WCDT-PCF (Fig. 3E-F) cohorts. As the *KMT2A* expression and MYC activity relationship become inverted upon progression to metastatic disease, the tight association with AR and HOXB13 lineage-specific drivers is presumably lost, suggesting a context-dependent role for KMT2A in PCa with increased MYC activity; increased *KMT2A* expression (and likely its activity) may drive development of primary disease but not advanced PCa.

**Figure 3.**
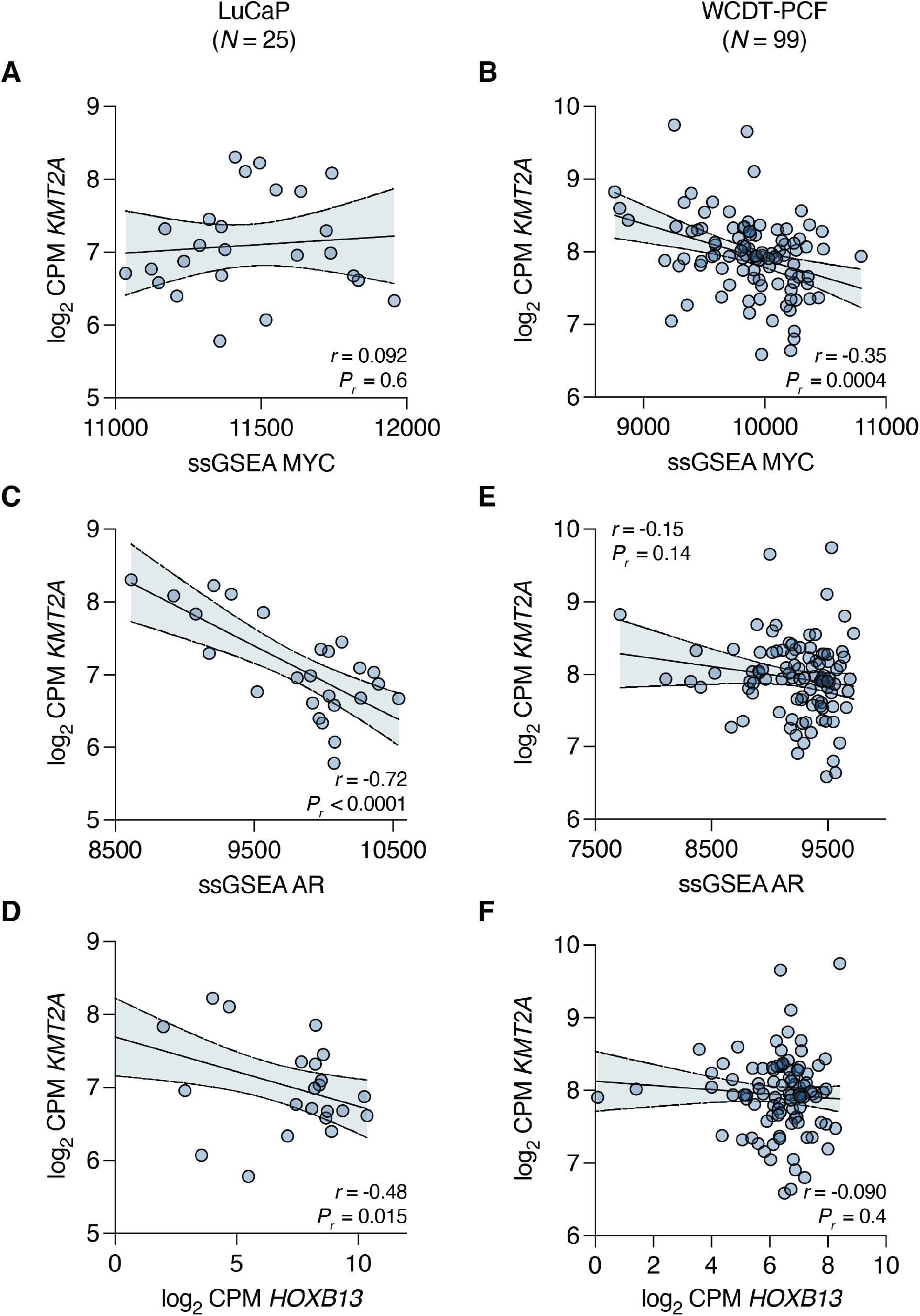
Inverse association of *KMT2A* expression with prostate cancer drivers in metastatic disease. **A, B, C, D, E, F**. Pearson correlation of the log_2_ CPM expression level for *KMT2A* with the 54-gene ssGSEA MYC activity score (A, B), a 266-gene ssGSEA AR activity score (C, E), or the log_2_ CPM expression level for *HOXB13* (D, F) in the LuCaP series of patient-derived xenografts (*N* = 25) (A, C, D) or the West Coast Dream Team-Prostate Cancer Foundation metastatic CRPC cohort (*N* = 99) (B, E, F). The error bars represent the 95% confidence bands for linear regression.

### Fluorine-18 labeled arachidonic acid uptake is increased in CRPC with low KMT2A expression and high MYC activity

Because *KMT2A* was differentially expressed relative to MYC activity in primary PCa versus mCRPC, we next examined potential functional implications of this relationship using the LuCaP PDX cohorts as a model of advanced disease in a series of ChIP-seq experiments. As depicted in Figure 4A, we stratified this cohort based on mean *KMT2A* expression (log_2_ CPM) across the entire LuCaP series, with ChIP-seq data available for 15 of these LuCaP models; nine were classified *KMT2A*-high and six were classified *KMT2A*-low. From ChIP-seq experiments with antibodies against AR and HOXB13, peaks were merged within each subgroup to create a union set of sites for each factor based on *KMT2A* status. For AR and HOXB13, we identified 180 and 203 differentially enriched peaks (cutoff of *P* < 0.1), respectively (Supplementary Table 2); the majority of which enriched in *KMT2A*-low samples (Fig. 4B). This enrichment is consistent with increased MYC-associated transcriptional activity of AR and HOXB13 in tumors with lower *KMT2A* expression (see Fig. 3B).

**Figure 4.**
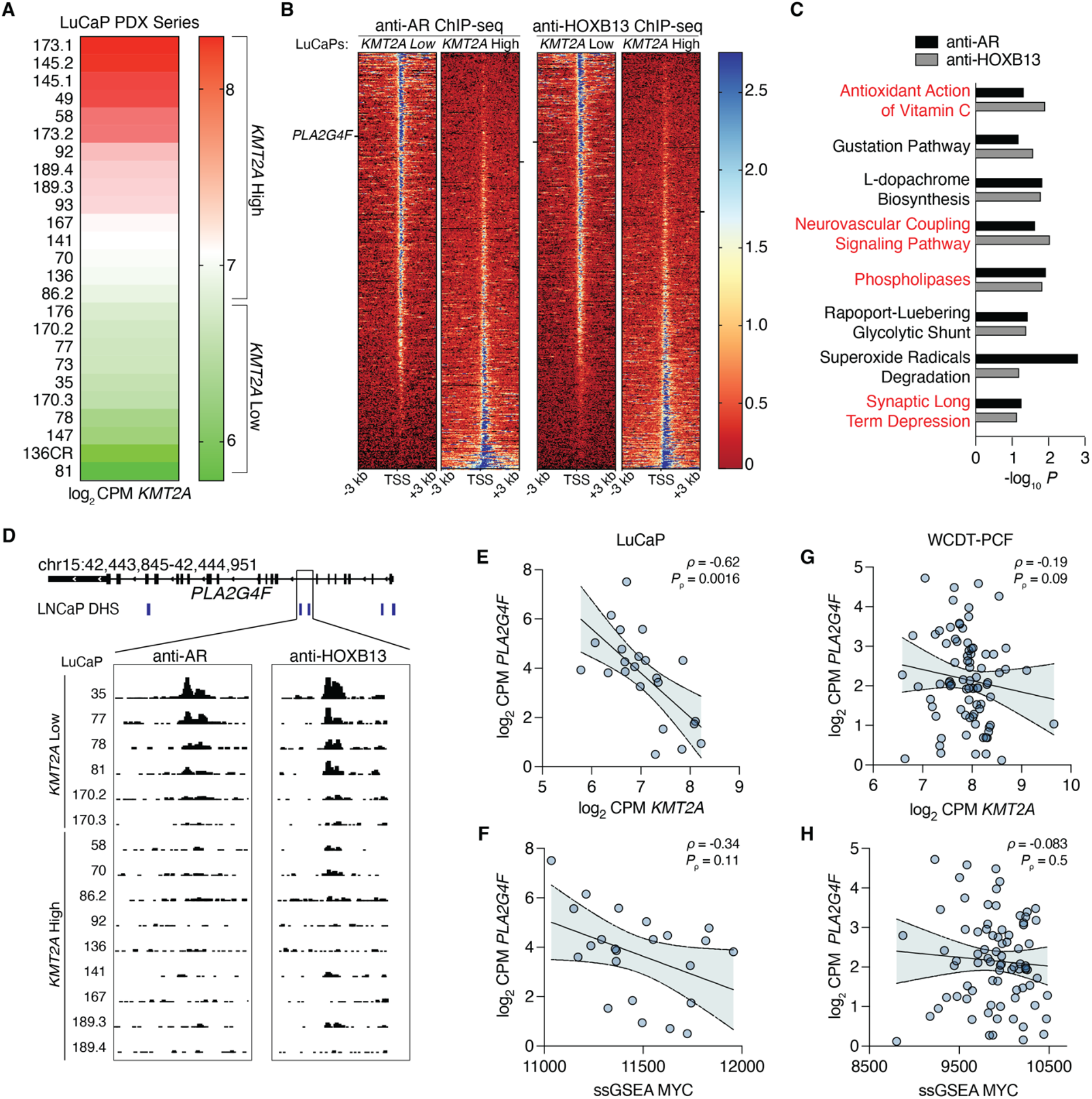
Inverse relationship between *PLA2G4F* and *KMT2A* expression in metastatic prostate cancer. **A**. Expression of *KMT2A* across the LuCaP PDX models in our study displayed as a heatmap, indicating which models are classified as *KMT2A* high or low. **B**. Heatmap of anti-AR and anti-HOXB13 ChIP-seq [29] peak signals (aggregated across samples as indicated) around TSS regions (± 3kb) in LuCaP PDX’s with differential *KMT2A* expression. Each row is a peak ranked by *KMT2A* low to high. The location of *PLA2G4F* on heatmap is marked by a black bar. **C**. Functional enrichment from Ingenuity Upstream Pathway Analysis of anti-AR and anti-HOXB13 ChIP-seq differential peaks overlapping genes for pathways with *P* < 0.1. Red indicates that *PLA2G4F is* present in the pathway gene set. **D**. Integrative genome viewer tracks of the *PLA2G4F* gene locus from anti-AR and anti-HOXB13 ChIP-seq. LNCaP DNase-hypersensitivity regions shown in blue. **E, F, G, H**. Spearman correlation of the log_2_ CPM expression level of *PLA2G4F* with log_2_ CPM expression level of *KMT2A* (E, G) or the 54-gene ssGSEA MYC activity score (F, H) in the LuCaP (E, F) or the West Coast Dream Team-Prostate Cancer Foundation metastatic CRPC cohort (G, H). Samples with log_2_ CPM *PLA2G4F <* 0 were excluded. The error bars represent the 95% confidence bands for linear regression.

We then sought to identify potential phenotypic effects in the *KMT2A*-low setting. We generated a list of differentially enriched genes for each transcription factor (TF) and histone mark and used these lists as input for IPA. IPA identified eight shared pathways, and examination of genes giving rise to pathway enrichment revealed strong representation of genes involved in fatty acid synthesis (Fig. 4C). The most common shared gene appearing in four of the eight pathways from both ChIP series, was *PLA2G4F* (phospholipase A2 group IVF), a key component of the arachidonic acid (ArA) metabolic pathway [33]. However, this enrichment was inversely proportional to *KMT2A* levels, with AR and HOXB13 being recruited to the *PLA2G4F* locus in the six *KMT2A*-low LuCaPs and mostly absent from the *PLA2G4F* locus in the nine *KMT2A*-high LuCaP models (Fig. 4D). Consistent with AR and HOXB13 binding, *PLA2G4F* mRNA expression was negatively correlated with *KMT2A* expression across all LuCaP PDXs (Fig. 4E) and maintained a negative correlation with MYC activity as well (Fig. 4F). This trend was also maintained in the WCDT-PCF mCRPC cohort (Fig. 4G-H). Interestingly, both *KMT2A* expression and MYC activity were positively correlated with *PLA2G4F* expression in primary PCa, with decreasing association stepwise from Gp3 to Gp4 to G5 (Supplementary Fig. 3). Taken together, these results further indicate a disparate role for *KMT2A* in MYC-driven advanced vs. localized PCa.

Due to the involvement of the protein product of *PLA2G4F*, cytosolic phospholipase A2 zeta, in ArA metabolism, we hypothesized that MYC activity*-*high, *KMTA2*-low, and *PLA2G4F*-high tumors have an increased cellular demand for ArA, due in part to its presence in phospholipids. We assessed uptake of fluorine-18 labeled ArA ([^18^F]ArA) over a two-hour time course in LuCaP PDX’s grown as organoids. As presented in Figure 5A, the rate of [^18^F]ArA uptake varied across LuCaPs, ranging from 6-88% per million cells in *KMT2A*-high LuCaP models and 27-154% per million cells in the *KMT2A*-low LuCaP models. Overall, *KMT2A*-low models demonstrated greater median [^18^F]ArA uptake at two hours (54%) than the *KMT2A*-high models (22%) (*P* = 0.026, Mann-Whitney *U* test; Fig. 5B). These findings indicate that differential co-regulation of *KMT2A* expression by the chromatin binding activities of MYC, AR, and HOXB13 is modulated during PCa progression and is associated with a shift in cellular metabolic requirements.

**Figure 5.**
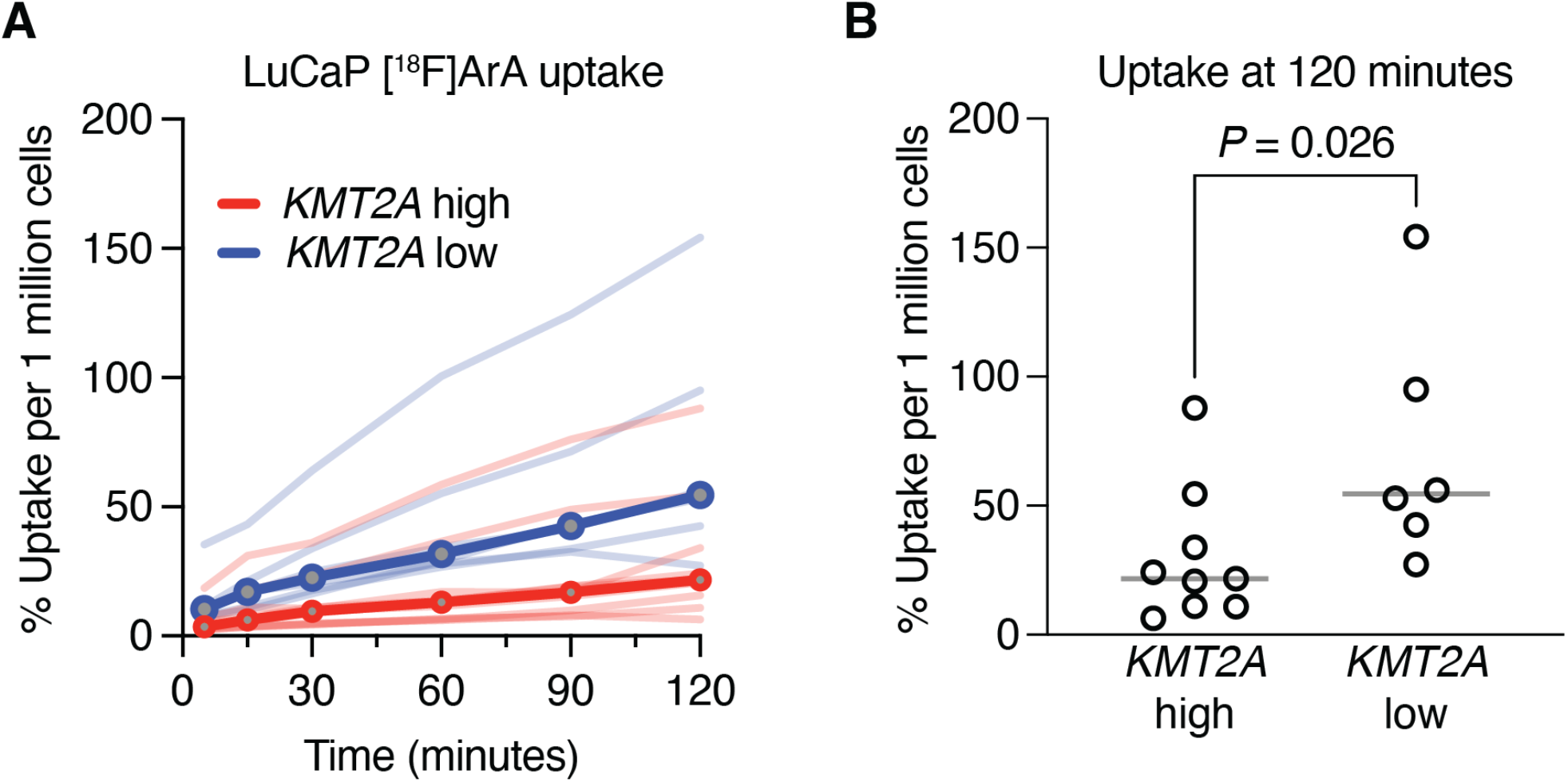
Arachidonic acid (ArA) metabolism is associated with *KMT2A* expression. **A**. Time course of [^18^F]ArA uptake in LuCaP PDX organoids (*N* = 15) classified as either *KMT2A-*high (red) or *KMT2A*-low (blue). Individual organoids are given by light shaded lines, with each line representing the median uptake of 2-5 replicates. Dark shaded lines represent the median uptake for *KMT2A-*high or *KMT2A*-low PDX organoids. Results are expressed as median uptake per 1 million cells. B. Percent [^18^F]ArA uptake in LuCaPs based on *KMT2A* expression at 120 minutes. Line at median. Each organoid (representing 2-5 replicate experiments) plotted as open circles (*N* = 9 high, *N* = 6 low). *P* = 0.026 by Mann-Whitney *U* test.

## DISCUSSION

Although compounds targeting MYC continue to be developed, many efforts to dissect and target the tumorigenic properties of MYC in PCa have shifted to its effector genes or co-factors necessary for its activity [34]. Thus, in this study, we started by assessing sites differentially bound by MYC, AR, and HOXB13 in the setting of high or low *MYC* expression using LNCaP PCa cells to identify novel transcriptional regulators that likely contributed to changes in MYC activity over the course of human PCa progression. From that analysis we identified *KMT2A/MLL1*, whose expression in association with MYC activity shifted from directly to inversely proportional as PCa progressed from localized to metastatic. Integrating ChIP-seq and WTS data from LuCaP PDX models revealed that AR and HOXB13 direct the regulation of lipid metabolism as a function of *KMT2A* levels.

The consequence of this finding, that MYC indirectly, but positively, regulates lipid metabolism in part through *PLA2G4F*, agrees with numerous reports that dysregulated *MYC* function drives glycolysis and lipogenesis, such that inhibition of MYC activity reduced accumulation of intracellular lipid droplets [17, 35]. The implications of increased lipid metabolism are two-fold: increased synthesis and turnover of membrane lipids associated with cell division, and fatty acid oxidation as an alternative energy source [36]. With respect to PCa specifically, studies have linked changes in ArA metabolism to PCa development and progression *in vitro*, with these studies suggesting that ArA is preferentially metabolized by PCa cells [37, 38]. However, to the best of our knowledge, the current study is the first to demonstrate an epigenetic link between ArA metabolism, uptake, and MYC activity in human prostate tumors.

The central premise of our finding relies on the observation that MYC is rewired from opposing *PLA2G4F* expression in early cancer to being positively associated with it in progressively advanced disease. The stepwise histologic progression from lower to higher Gleason grades is reflective of dedifferentiation, which in turn is a direct consequence of altered AR activity [39]. Similarly, AR-directed therapies suppress AR activity in patients with metastatic PCa, such that resistant tumors are less dependent on AR for growth and survival [39]. Thus, histologically and clinically aggressive tumors may reflect a causal link between increased MYC activity and decreased but persistent AR activity, due in part to a transcriptional pause-release effect that accounts for lower AR tumors having worse clinical outcomes [4, 40]. A logical future study will be to broadly assess the metabolic and lipogenic ramifications of differential AR/MYC status.

This current study has two important limitations. First, we observed a switch in the association between *KMT2A* or *PLA2G4F* expression and MYC activity, AR activity, and *HOXB13* expression transcriptomically between primary localized PCa. Although we could recapitulate the inverse association using ChIP-seq from LuCaP PDX models for which matched transcriptional data was available, similar ChIP-seq assays from primary PCa with matched RNA-seq were not obtainable. Second, we demonstrated functional changes in lipid metabolism using [^18^F]ArA uptake as a surrogate for cellular metabolic needs. Measurements of actual cPLA_2_ protein levels would have been ideal, but after extensive testing, none of the four commercial antibodies with described reactivity against the Group IV *zeta* (*PLA2G4F*) isoform were specific.

Despite these limitations, we have demonstrated concordance between transcriptional and chromatin readouts of TF activity within the same panel of PDX models, with further qualification of potential lipid metabolic differences using the PDXs in an *in vitro* functional assay. Our findings indicate that ArA metabolism is specifically sensitive to AR and MYC activity during PCa progression.

## CONCLUSIONS

We have reported that ArA metabolism and uptake are enriched in MYC-high prostate tumors. This finding has implications for the subset of treatment resistant prostate cancers that maintain high MYC activity and develop resistance to both hormonal and cytotoxic chemotherapies. Future studies are needed to validate these findings in larger cohorts, assess the viability of targeting ArA metabolism therapeutically, and determine whether ArA-based imaging agents demonstrate specific uptake in MYC-high/*KMT2A*-low prostate cancers.

## METHODS

### Study design

The aim of this study was to assess changes in gene expression patterns and corresponding cellular phenotypes associated with repositioning of MYC in prostate cancer. This study utilized LNCaP cells engineered to have reduced levels of *MYC* expression as described in [22], LuCaP PDX organoids as described in [31], and integration of both novel and published datasets derived from these lines and patient cohorts [22, 23, 29–31, 41].

### ChIP-seq analysis in LNCaP cells

ChIP-seq against MYC in LNCaP cells harboring a nontargeting control hairpin (NTC) or *MYC*-targeting hairpins (*MYC* shRNA) was described previously [22]. ChIP-seq performed in LNCaP cells or LuCaP patient-derived xenografts (PDXs) were previously described [23, 29] and were retrieved from GEO (https://www.ncbi.nlm.nih.gov/geo/) using accession numbers GSE94682 and GSE130408, respectively. Downloaded FASTQ files were reprocessed through the nextflow [42] nf-core/chipseq (v1.2.2) pipeline (https://github.com/nf-core/chipseq). De-multiplexed reads were aligned to build GRCh37 of the human genome using BWA-MEM [43]. MACS2 [44] was used to perform peak calling using a cutoff FDR *q*-value of 0.01 and the parameter --narrow_peak. HOMER [45] annotePeaks.pl was used to annotate called peaks relative to known genomic features.

### RNA-seq analysis

Whole transcriptome profiling of laser capture microdissected (LCM) primary prostate cancer (PCa) was previously described [22]. Transcriptomes from the Stand Up To Cancer West Coast Dream Team-Prostate Cancer Foundation metastatic castration resistant prostate cancer (WCDT-PCF mCRPC) cohort [41] were downloaded from the NCI Genomic Data Commons (https://gdc.cancer.gov) via access to dbGaP phs001648. Transcriptomes from the LuCaP [23, 29–31] series of patient-derived xenografts (PDXs) were downloaded from GEO using accession numbers GSE113741, GSE156292, and GSE126078. Downloaded FASTQ files were processed through the nextflow nf-core/rnaseq (v3.5) pipeline (https://github.com/nf-core/rnaseq). Sample reads were mapped to the GRCh37 reference genome using STAR [46] and quantified using featureCounts [47] with default parameters. EdgeR (v3.32.1) was used to generate the log_2_ counts per million (CPM) values used for downstream analysis [48].

Single-sample gene set enrichment (ssGSEA) was performed using the R package GSVA (v1.38.2) with the following parameters: tau: 0.75; ssgsea.norm: FALSE [49]. For each sample, ssGSEA projection values were obtained for a 54-gene MYC activity score [50] or a 266-gene AR activity score [51].

### Identification of core and redistributed ChIP-seq peaks in LNCaP cells

Bed files for LNCaP ChIP-seq peaks created by MACS2 were used as input for BEDTools (v2.30.0) [52] to identify *core* and *redistributed* peaks. Each independent MYC ChIP-seq (NTC and *MYC* shRNA) was compared to AR or HOXB13 ChIP-seq data. Here, *core* (overlapping) peaks were defined as co-occupied binding sites enriched in both NTC- and *MYC* shRNA-harboring LNCaP cells. *Redistributed* (unique) peaks were defined as co-occupied binding sites enriched in *MYC* shRNA cells. New peak files representing this intersection were collated where appropriate and annotated to the nearest gene body (GB) or transcription start site (TSS) within ±3kb, and redundant gene symbols were removed.

### ChIP-seq analysis in LuCaP xenografts

LuCaP xenografts for which ChIP-seq data were available were classified as *KMT2A*-high (*N* = 9) or *KMT2A*-low (*N* = 6) based on corresponding average *KMT2A* log_2_ CPM expression values across the entire LuCaP cohort, including cases where ChIP-seq was not performed. DESeq2 (v1.30.1) was used to identify differential peaks between *KMT2A*-high and *KMT2A*-low samples selecting peaks with adjusted *P* < 0.05 [53]. BigWig files were merged to create a consensus wiggle file based on *KMT2A* status for each ChIP-seq using WiggleTools (v1.2) [54] and converted into bigWig files using the UCSC wigToBigWig tool [55]. Generated bigWig files were visualized with the integrative genomics viewer. ChIP-seq heatmaps were prepared using deepTools (v3.5.1) computeMatrix and plotHeatmap tools [56].

### Pathway Analysis

Core analyses were performed with Ingenuity Pathway Analysis (IPA) tools [57]. For deduplicated *core* and *redistributed* peak lists derived from LNCaP ChIP-seq data, Upstream Regulator Analysis (URA) was used to identify factors upstream of the genes in the peak lists. URA was limited to *transcription regulators*. Enriched gene sets selected by GSEA (FDR *q* < 0.05) were used as input for Canonical Pathway analysis and normalized gene counts for shared core enrichment genes identified for primary PCa or CRPC were used. Counts were normalized as follows: (*MYC* counts + *KMT2A* counts)/total shared gene sets. Consensus differential peaks for LuCaP ChIP-seq data were also used for Canonical Pathways analysis, with overlapping pathways selected at a cutoff of *P* < 0.1.

### Fluorine-18 labeled Arachidonic acid ([^18^F]ArA) uptake in LuCaP xenografts

LuCaP xenografts were processed and cultured as organoids as described previously [31]. Organoid identity was validated every six months by STR profiling (Laragen). Organoids were cultured for 7 days in 2% Matrigel, collected, washed with PBS, then resuspended in 1 mL of Beshiri’s Modified Clevers Media [31] in duplicate tubes for *in vitro* [^18^F]ArA uptake assays.

[^18^F]ArA was prepared following as previously described with minor modifications [58]. Briefly, the tosylate precursor was heated with [^18^F]TBAF in acetonitrile at 120 °C for 20 min followed by hydrolysis with potassium hydroxide (120 °C for 10 min) to produce 20-[^18^F]ArA. The overall radiochemical yields were 15-24% (uncorrected, *n* > 6) with a molar activity of 59-163 GBq/μmol (n > 6).

Organoid cultures were radiolabeled with 2μCi [^18^F]ArA in BMCM for 120 minutes at 37°C/5% CO_2_, washed twice with PBS to remove residual unincorporated [^18^F]ArA, and resuspended in 1mL trypsin as a single-cell suspension. Cells were counted using acridine orange/propidium iodide cell counting dye. [^18^F]ArA uptake was determined by PerkinElmer 2480 Wizard3 Gamma Counter. For each sample, percent uptake per 10^6^ live cells was calculated as follows:

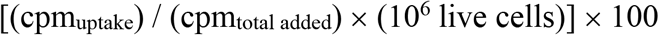

### Statistical analysis

Statistical analyses were performed with GraphPad Prism version 9 (GraphPad Software) for Mac. Associations between factors were measured using Pearson or Spearman correlations. Comparisons of single factors between dichotomized samples was performed using Mann-Whitney *U* tests.

## Data Availability

The data underlying this article have been deposited in Database of Genotypes and Phenotypes (dbGaP) and Gene Expression Omnibus (GEO) at https://www.ncbi.nlm.nih.gov/gap/ and https://www.ncbi.nlm.nih.gov/geo/, respectively, and can be accessed with phs001813.v2.p1 (dbGaP), phs001938.v3.p1 (dbGaP) and GSE135942 (GEO).

https://www.ncbi.nlm.nih.gov/gap/

## Acknowledgements

The authors gratefully acknowledge the patients and the families of patients who contributed to this study. Portions of this work utilized the computational resources of the NIH HPC Biowulf cluster.

## LIST OF ABBREVIATIONS

[^18^F]ArA: Fluorine-18 labeled arachidonic acid
AR: Androgen receptor
ArA: Arachidonic acid
CRPC: Castration-resistant prostate cancer
mCRPC: Metastatic castration-resistant prostate cancer
PCa: Prostate cancer
PDX: Patient-derived xenograft
TF: Transcription factor

