## Supplementary Information for "Progression of prostate cancer reprograms MYC-mediated lipid metabolism via lysine methyltransferase 2A"

by

Nichelle C. Whitlock et al.

Supplementary Methods
Supplementary Table Legends
Supplementary Figures
Supplementary References

### Supplementary Methods

*Microarray analysis of laser capture microdissected prostate tumor foci*

Affymetrix Human Exon Array data of 26 laser capture microdissected human prostate tumors of Gleason patterns 3 and 4 (1) were downloaded from GEO using accession number GSE52560. SCAN.UPC (2) normalized intensity values were used.

*RNA-seq analysis of TCGA tumors*

Whole transcriptomes of The Cancer Genome Atlas prostate adenocarcinoma (TCGA-PRAD) primary prostate cancer cohort (3) were retrieved from the NCI Genomics Data Commons (https://gdc.cancer.gov) via access to dbGaP phs000178. Cases with Gleason scores of 3+5=8 and 5+3+8 were excluded. Sample reads were aligned and processed as with the primary LCM RNA-seq cohort.

### Supplementary Table Legends

**Supplementary Table 1.** Ingenuity upstream regulator candidates based on gene body or transcriptional start site loci for anti-AR or anti-HOXB13 ChIP-seq. For each potential upstream regulator, its rank from either core or redistributed sites is shown along with the *P* value of overlap with the canonical IPA gene set for that regulator.

**Supplementary Table 2**. Differentially enriched genes corresponding to AR- or HOXB13-bound sites (peaks) based on *KMT2A* expression.

### Supplementary Figures

#### Supplementary Figure 1

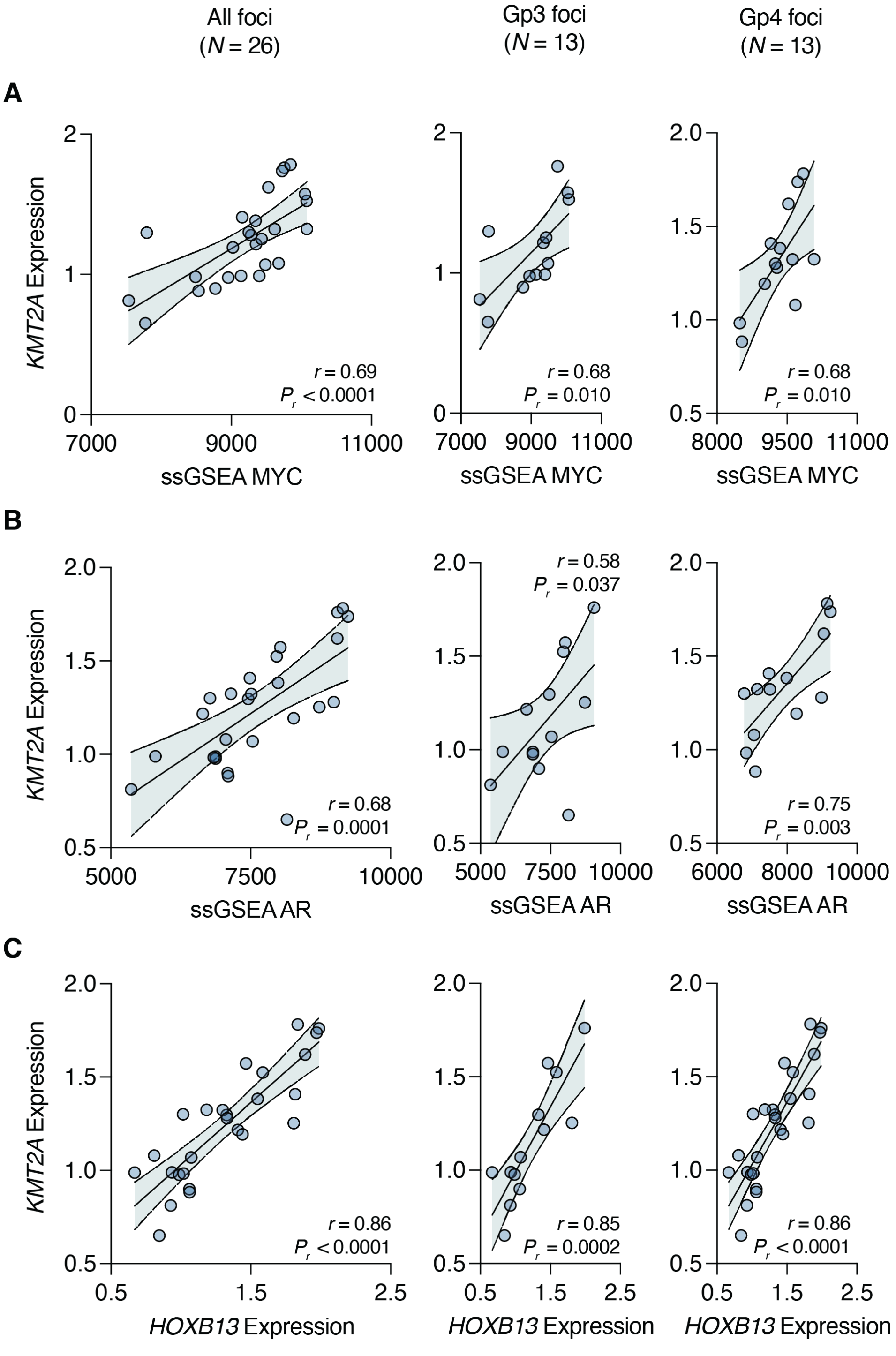


**Supplementary Figure 1. Association of *KMT2A* expression with prostate cancer drivers in primary disease. A, B, C.** Pearson correlation of the normalized expression of *KMT2A* with a 54-gene ssGSEA MYC activity score (A), a 266-gene ssGSEA AR activity score (B), or the normalized expression of *HOXB13* (C) a cohort of 26 laser capture microdissected foci of human prostate tumors). The error bars represent the 95% confidence bands for linear regression. For each correlation, the foci are subdivided into Gp3 (left), Gp4 (right, including intraductal tumors).

### Supplementary Figure 2

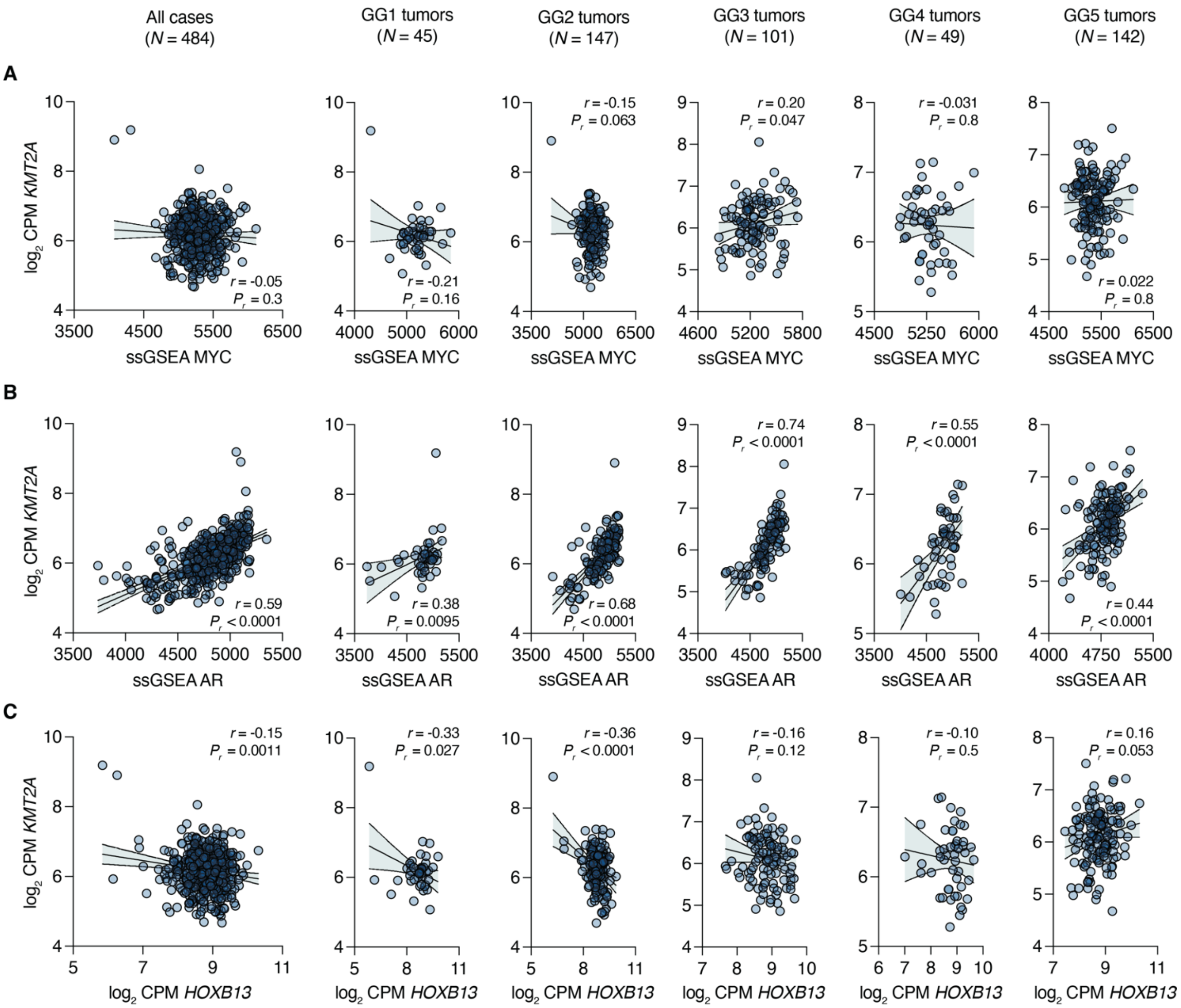


**Supplementary Figure 2. Association of *KMT2A* expression with prostate cancer drivers in primary disease. A, B, C.** Pearson correlation of the $\log_2$ CPM expression level for *KMT2A* with a 54-gene ssGSEA MYC activity score (A), a 266-gene ssGSEA AR activity score (B), or the $\log_2$ CPM expression level for *HOXB13* (C) a cohort of 484 human prostate tumors from the prostate cancer TCGA (TCGA-PRAD). The error bars represent the 95% confidence bands for linear regression. For each correlation, the foci are subdivided into Gleason ISUP Grade Groupings (1-5); tumors scored Gleason 3+5=8 or 5+3=8 excluded.

**Supplementary Figure 3**

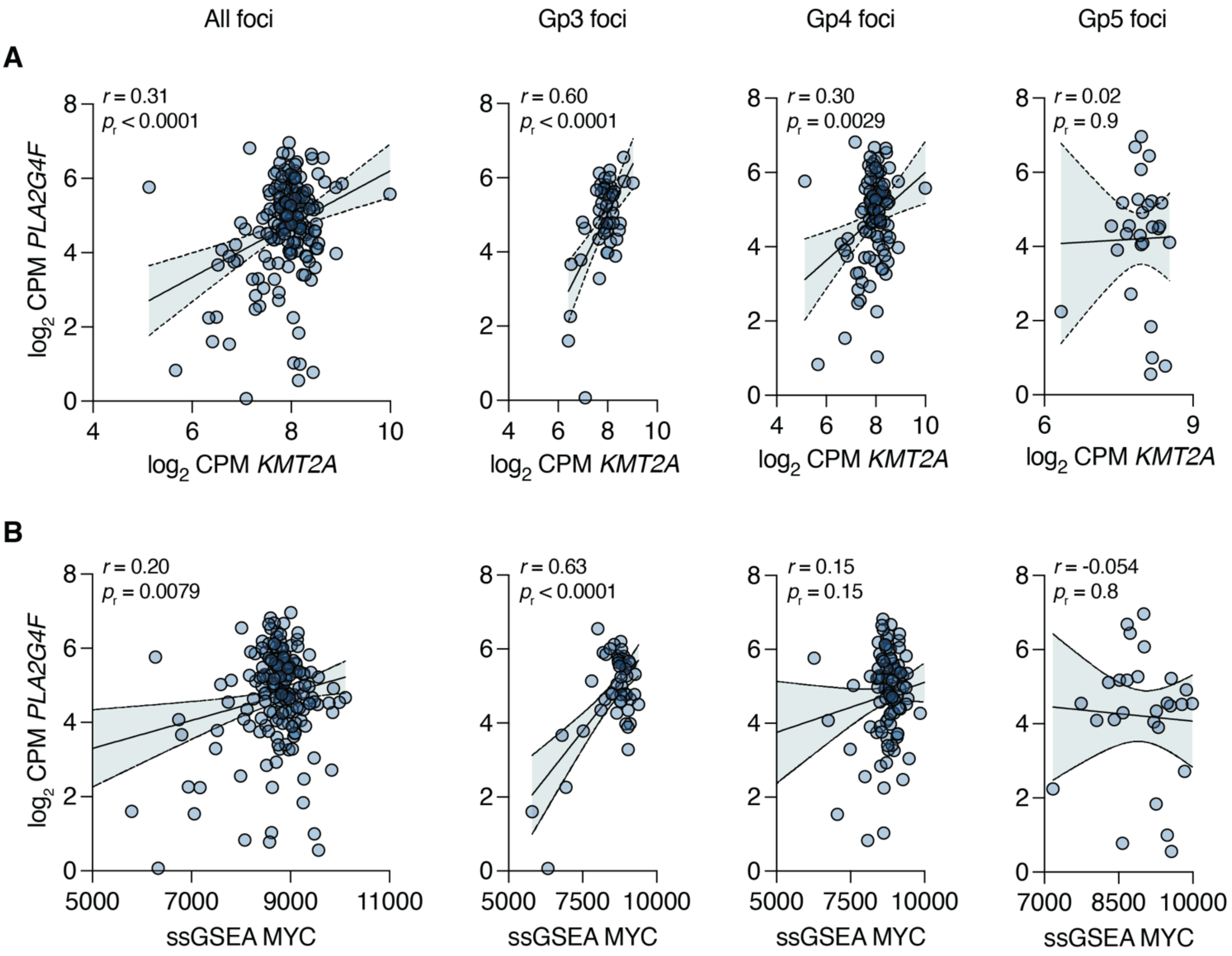


**Supplementary Figure 3. Decreasing association between *PLA2G4F* and *KMT2A* expression and MYC activity in primary prostate cancer. A, B.** Pearson correlation of the $\log_2$ CPM expression level of *PLA2G4F* with $\log_2$ CPM expression level of *KMT2A* (A) or the 54-gene ssGSEA MYC activity score (B) in a cohort of laser capture microdissected foci of human prostate tumors. The error bars represent the 95% confidence bands for linear regression. Samples with $\log_2$ CPM *PLA2G4F* < 0 were excluded. For each correlation, the foci are subdivided into Gp3 (left), Gp4 (middle, including intraductal tumors), and Gp5 (right).
